# Rural-Urban Disparities of Alzheimer’s Disease and Related Dementias: A Scoping Review

**DOI:** 10.1101/2024.11.08.24316977

**Authors:** Mackenzie Kramer, Maxwell Cutty, Sara Knox, Alexander V. Alekseyenko, Abolfazl Mollalo

## Abstract

**Background & Aim:** The rising age of the global population has made Alzheimer’s Disease and Related Dementias (ADRD) a critical public health problem, with significant health-related disparities observed between rural and urban areas. However, no previous reviews have examined the scope and determinant factors contributing to rural-urban disparities of ADRD-related health outcomes. This study aims to systematically collate and synthesize peer-reviewed articles on rural-urban disparities in ADRD, identifying key determinants and research gaps to guide future research.

**Methods:** We conducted a systematic search using key terms related to rural-urban disparities and ADRD without restrictions on geography or study design. Five search engines—MEDLINE, CINAHL, Web of Science, PubMed, and Scopus—were utilized to identify relevant articles. The search was performed on August 16, 2024, and included articles published from 2000 onward.

**Results:** 62 articles met the eligibility criteria for data extraction and synthesis. Most articles were published after 2010 (90.3%) and were concentrated in the US, China, and Canada (64.5%). A majority had cross-sectional (59.7%) or cohort study designs (24.2%), primarily examining prevalence (40.3%) or incidence (11.3%). Findings often indicated a higher prevalence and incidence in rural areas, although inconsistent rural-urban classification systems were noted. Common risk factors included female gender, lower education level, lower income, and comorbidities such as diabetes and cerebrovascular diseases. Environmental (12.9%) and lifestyle (14.5%) factors for ADRD have been less explored. The statistical methods used were mainly traditional analyses (e.g., logistic regression) and lacked advanced techniques such as machine learning or causal inference methods.

**Conclusion:** The gaps identified in this review emphasize the need for future research in underexplored geographic regions and encourage the use of advanced methods to investigate understudied factors, such as environmental, lifestyle, and genetic influences.

## 1. INTRODUCTION

As the global population of older adults surges, Alzheimer’s Disease and Related Dementias (ADRD) have become an urgent public health problem [1, 2]. Although ADRD affects people across a wide geographic range, its impact—and the efforts and resources available to address it—vary significantly between rural and urban areas [3, 4]. A 2021 data brief from the Centers for Disease Control and Prevention indicated that among the ten leading causes of death in 2019, mortality rates were higher in rural areas than in urban areas, with Alzheimer’s disease ranking as the sixth leading cause of death [3]. From 1999 to 2019, rural areas experienced a rapid increase in ADRD mortality, significantly contributing to widening life expectancy gaps compared to urban regions [5].

More specifically, the existing research has shown that rural-urban disparities in ADRD largely result from the cumulative stress and strain of lifelong socioeconomic challenges that disproportionately impact rural populations [6]. These challenges arise from a combination of risk factors (e.g., low education, social isolation, chronic diseases), population dynamics (e.g., depopulation, racial inequality), and fragmented healthcare systems, all of which adversely affect access to, utilization of, and quality of care in rural areas [7, 8]. Consequently, it is unsurprising that most research suggests that rural populations have a higher risk of ADRD and that rural healthcare systems are less well-equipped to provide quality care tailored to the needs of people with ADRD.

While sustained public health and health policy efforts have made significant strides in reducing rural-urban disparities in health outcomes of ADRD, much of the research informing these initiatives has focused on a few developed countries, as well as simplistic dementia classification systems, traditional methodological approaches (e.g., logistic regression), and limited outcome measures [9, 10]. This has resulted in significant gaps in the evidence base, underscoring the need for a comprehensive review of the literature on ADRD disparities between rural and urban areas. To address the gaps, this scoping review seeks to provide a descriptive overview of previous peer-reviewed studies with the following objectives:

1. to highlight geographic areas where further research is needed to enhance the global understanding of rural-urban disparities of ADRD-related health outcomes;
2. to describe different rural-urban classification systems used in studies of ADRD-related health outcomes;
3. to analyze differences in health outcomes of ADRD (e.g., incidence, prevalence, mortality, healthcare utilization) and access, utilization and quality of care between rural and urban settings;
4. to identify factors (e.g., environmental, socioeconomic, lifestyle, comorbidities) that have been associated with rural-urban disparities of ADRD-related health outcomes;
5. to describe statistical methods used in examining rural-urban disparities of ADRD-related health outcomes.

The findings of this study aim to encourage researchers to explore the gaps and foster innovative approaches for future investigations.

## 2. METHODS

We adhered to the Preferred Reporting Items for Systematic Reviews and Meta-Analyses extension for Scoping Reviews (PRISMA-ScR) guidelines throughout our review process to identify the articles that satisfy the eligibility criteria for final inclusion in this scoping review. The PRISMA checklist is provided in Appendix 1.

### 2.1. Data sources

A comprehensive search strategy for peer-reviewed articles was implemented, involving thorough screenings of the abstracts and titles from five search engines: MEDLINE, CINAHL, Web of Science, PubMed, and Scopus.

### 2.2. Search strategy

The search, performed on August 16, 2024, applied no restrictions on study designs or geography. Our initial screening of abstracts and titles included all peer-reviewed, English-language articles that contained key terms related to rural, urban, and ADRD. We used logical operators to combine terms related to ‘rural’, ‘urban’, and ‘ADRD’ themes. Table 1 presents the complete list of key terms. We employed the advanced search options on each database to identify the articles that included these terms.

**Table 1.** Search key terms.

| Theme | Key Terms |
| --- | --- |
| Rural | ("rural" OR "ruralities" OR "rurality" OR "rurally" OR "ruralness")<br>AND |
| Urban | ("urban" OR "urbanicity" OR "urbanism" OR "urbanity" OR "urbanization" OR<br>"urbanizations" OR "urbanize" OR "urbanized" OR "urbanizes" OR<br>"urbanizing")<br>AND |
| ADRD | ("Dementia" OR "Alzheimer Disease" OR "Alzheimer*" OR "ADRD") |

### 2.3. Article selection

All abstracts and titles obtained from each database were imported into the Covidence online tool (https://www.covidence.org/ accessed on August 2, 2024). Duplicates were removed through automatic recognition by Covidence and manual identification by the reviewers. Two reviewers, MK and MC, independently screened the abstracts and titles, excluding articles where ADRD was not the main outcome. Moreover, we excluded reviews, case reports, editorials, symposiums, gray literature, and conference proceedings. Any conflicts during the abstract screening process were resolved through follow-up discussions until a consensus was reached. The same two reviewers screened the full texts of the selected articles. Articles were excluded during the full-text review for the following reasons: the primary outcome was not ADRD, the article was published before the year 2000, the full-text was not available, the article was not peer-reviewed, the article lacked rural-urban comparisons, or the article did not focus on ADRD patients. Again, conflicts during the full-text screening process were resolved through follow-up discussions until a consensus was achieved. In cases where full texts could not be accessed in any database, we contacted the authors directly to request copies of the articles.

Figure 1 depicts the article selection process.

**Figure 1.**
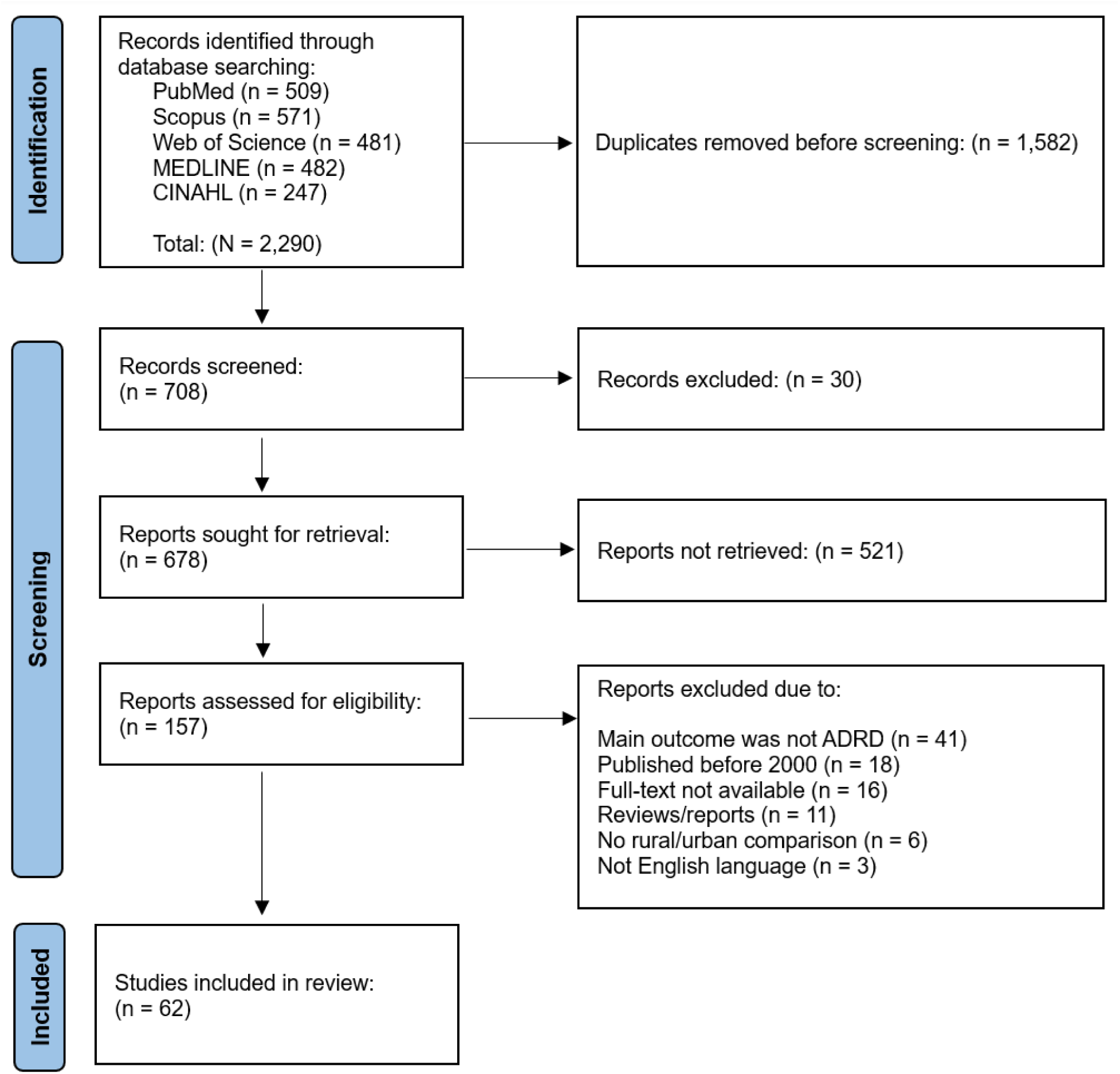
PRISMA flowchart for the article selection process.

### 2.4. Data extraction and synthesis

After identifying the articles that met all the eligibility criteria, we extracted data from each article. This included the article title, author names, location, year of publication, level of analysis (e.g., individual, county), study design (e.g., cohort, cross-sectional, ecological), outcome investigated (e.g., Alzheimer’s disease, vascular dementia), sample size, and rural-urban classification method. Moreover, we extracted various risk factors such as demographics, socioeconomics, environment, lifestyle, and comorbidities. Additional attributes included healthcare access and utilization, quality of care, interventions or policies (e.g., screening programs), statistical methods, and study limitations. Following data extraction, we provided a narrative synthesis by combining the article’s characteristics to identify key determinants contributing to the disparities and highlight significant research gaps that warrant further investigation.

## 3. RESULTS

### 3.1. Article selection

We initially identified 2,290 articles from the MEDLINE, CINAHL, Web of Science, PubMed, and Scopus databases. After removing 1,612 duplicates through automatic identification via Covidence and manual checks, we were left with 678 unique articles for screening. During the abstract and title screening, 521 articles were deemed irrelevant and removed. Full-text reviews were conducted on the remaining 157 articles, resulting in the removal of an additional 95 articles. Reasons for removal include dementia not being the primary outcome (n=41), articles published before the year 2000 (n=18), lacking full-text availability (n=16), being reviews/reports (n=11), not containing a rural-urban comparison (n=6), and being published in a language other than English (n=3). The final selection comprised 62 articles. Detailed characteristics of each article are provided in Appendix 2.

### 3.2 Geographic and temporal distribution of articles

Of the 62 articles selected, 35.5% (n=22) were conducted in the US [11–32], 22.6% (n=14) in China [33–46], 6.4% (n=4) in Canada [47–50], 4.8% (n=3) in Sweden [51–53], 3.2% (n=2) each in Germany [54, 55], Taiwan [56, 57], South Korea [58, 59], and India [60, 61], and 1.6% (n=1) each in Indonesia [62], Egypt [63], Japan [64], England [65], Spain [66], Bangladesh [67], Pakistan [68], Portugal [69], Central African Republic [70], and Great Britain [71]. Only 1.6% of articles (n=1) was a multi-country article, including China, India, Cuba, the Dominican Republic, Venezuela, Mexico, and Peru [72]. Figure 2A depicts the geospatial distribution of articles by country. Most articles (n=56, 90.3%) were published after 2010, particularly post-2017 (n=42, 67.7%). 2020 was the year with the highest number of publications (n=7, 11.3%), closely followed by 2018, 2021, 2022, and 2023 (n=6, 9.7% each year). Figure 2B shows the temporal trend of the publications included.

**Figure 2.**
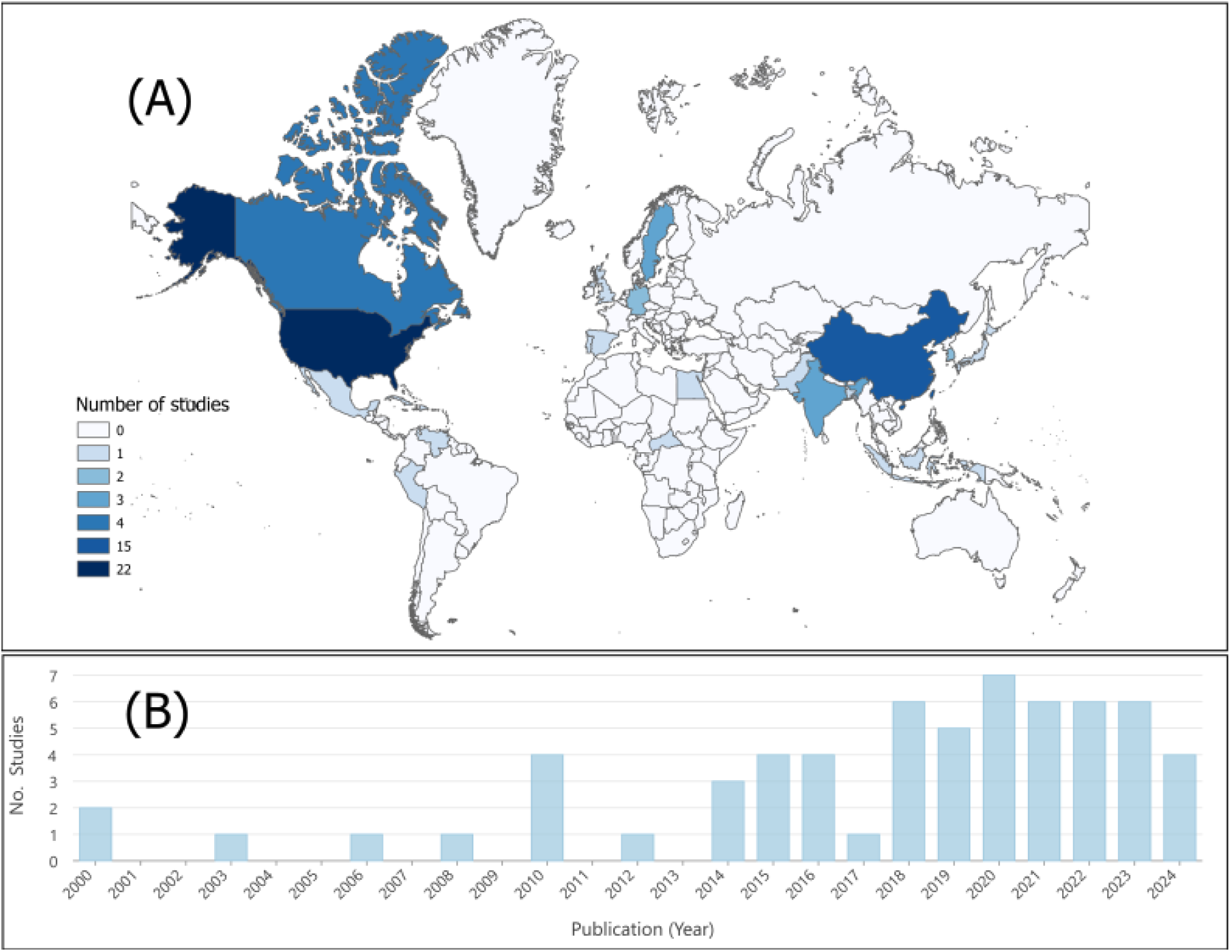
A) Geographical distribution and B) temporal distribution of the included articles.

### 3.3. Rural-urban classification systems

Rural-urban classification systems within each article varied at both inter- and intra-country levels across the US (n=22, 35.5%), China (n=14, 22.6%), Canada (n=4, 6.4%), Sweden (n=3, 4.8%), and other countries (n=19, 30.6%). Of the 22 articles performed within the US, several articles utilized Rural-Urban Continuum Codes (RUCC) (n=8, 36.4%), followed by Rural-Urban Commuting Area Codes (RUCA) (n=4, 18.2%), Urban Influence Codes (UIC) (n=2, 9.1%) [17, 28], and arbitrary classification systems based on population size (n=7, 31.8%). One article within the US did not specify its classification system [20]. Of the 14 articles from China, the majority did not specify their rural-urban classification (n=11, 78.6%), while the remaining articles used a variety of systems, including a binary classification (n=1, 7.1%) [40], jurisdictional designations (n=1, 7.1%) [37], and administrative characteristics (n=1, 7.1%) [45]. Four articles from Canada employed a variety of rural-urban classification methods, including Statistical Area Classification (n=1, 25%) [47], the second digit of postal codes (n=1, 25%) [48], defining rural areas as populations of 1,000 or fewer with a density of less than 400 people/km² (n=1, 25%) [49], and defining rural as populations below 10,000 (n=1, 25%) [50]. Other classification methods include defining rural as a density of fewer than 150 inhabitants/km² (n=1, 0.1.6%) [54], a population below 10,000 (n=1, 1.6%) [71], and a population below 300,000 individuals that is not adjacent to communities with populations of 300,000 individuals or more (n=1, 0.1.6%) [64]. Many of the remaining articles failed to provide a clear definition of rural (n=11, 57.9%).

### 3.4. Study designs and outcomes

The main outcomes investigated were prevalence (n=25, 40.3%), followed by healthcare utilization (n=8, 12.9%), incidence (n=7, 11.3%), mortality (n=5, 8.1%), places of care (n=4, 6.5%), diagnostics (n=3, 4.8%) [20, 32, 52], preventable emergency department visits (n=2, 3.2%) [29, 30], and age of diagnosis (n=1, 1.6%) [58]. There was only one article (n=1, 1.6%) for each of the following categories: treatment severity [41], years of life lost [33], quality of life [71], patient types among general practitioners (GPs) [54], neuropsychiatric symptoms [13], hospitalizations [28], and undetected prevalence among those with dementia [43] (Figure 3A).

**Figure 3.**
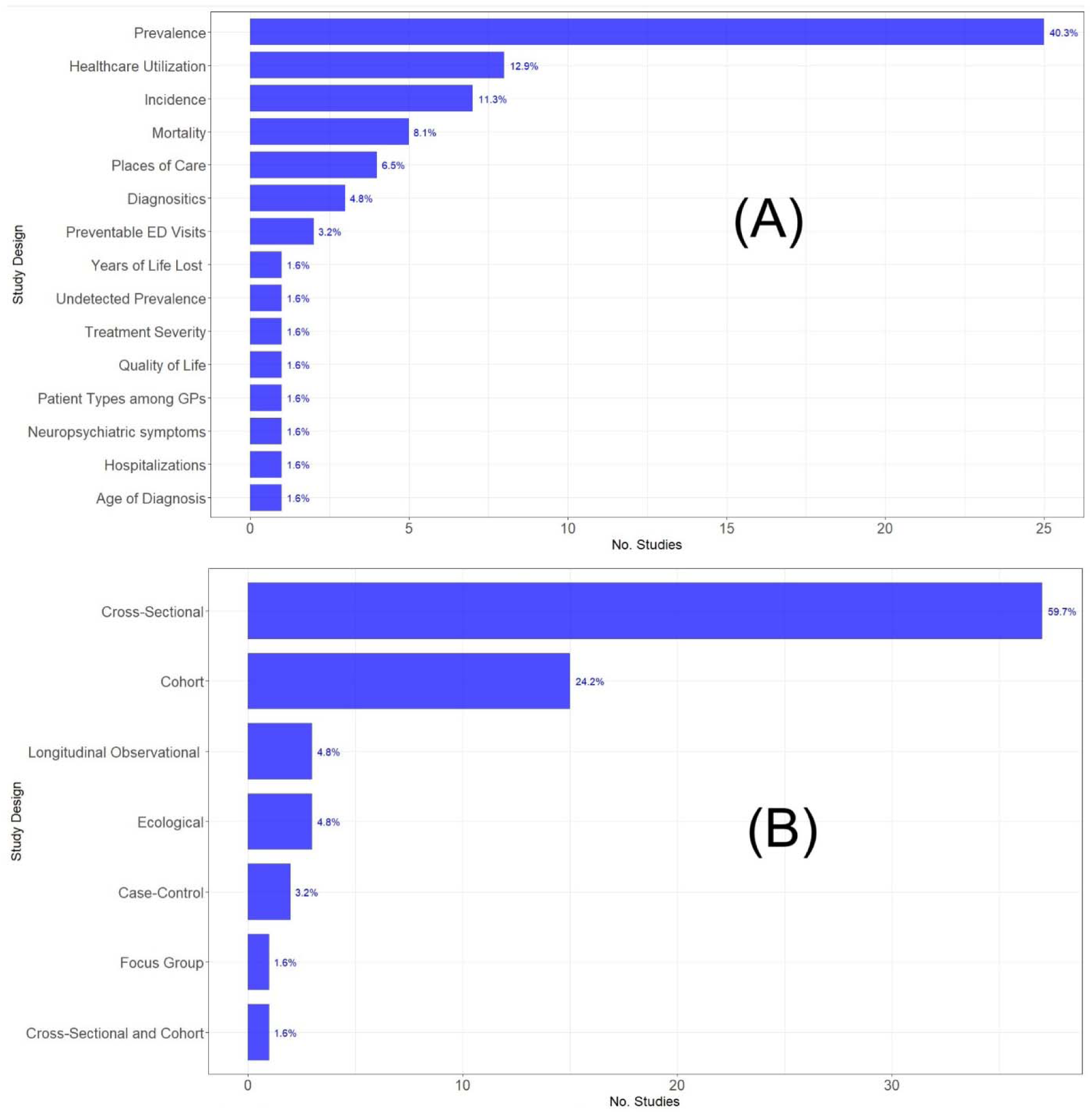
The distribution of (A) health outcomes and (B) study designs. ED: Emergency department; GPs: General practitioners.

The most frequent study designs were cross-sectional (n=37, 59.7%), followed by cohort (n=15, 24.2%). Other study designs included ecological (n=3, 4.8%) [11, 14, 24], longitudinal observational (n=3, 4.8%) [33, 42, 58], case-control (n=2, 3.2%) [55, 56], focus groups (n=1, 1.6%) [54], and a combined cross-sectional and cohort (n=1, 1.6%) [46] (Figure 3B). Most of the included articles were performed at the individual level (n=57, 92%), while a limited number of articles were conducted at the county level (n=4, 6.5%), and only one article (n=1, 1.6%) was performed at the surveillance site level [33]. Sample sizes varied greatly, with an average of slightly over 6.8 million. Stratified by study design, ecological articles typically had the smallest sample sizes, ranging from 8 surveillance sites to 175 counties (SD=71). Cohort articles had a range of 181 to 2.9 million participants (SD=1,240,861). Cross-sectional articles ranged from 146 to 328 million participants (SD=11,814,498).

Findings of disparities in the prevalence and incidence of ADRD between rural and urban areas differed across articles. Among the 29 articles that covered rural-urban differences in prevalence or incidence, most of them found a higher prevalence and incidence of ADRD in rural areas (n=17, 58.6%), but others found the opposite (n=5, 17.2%) or no statistically significant relationship (n=6, 20.7%). Furthermore, individuals from urban areas exhibited a higher prevalence of neuropsychiatric symptoms than rural areas (n=1, 3.6%) [13]. Nearly all articles investigated secondary and tertiary outcomes, which will be discussed more in the following sections.

The forms of dementia investigated varied across articles. Nearly half of the articles stated ‘dementia’ as their outcome of interest, with no further explanation of the type of dementia included (n=30, 48.4%). Other common types of dementia studied include ADRD (n=14, 22.6%), Alzheimer’s Disease (AD) specifically (n=9, 14.5%), and the remaining articles related to specific forms of dementia such as AD, vascular dementia, or Parkinson’s Disease Dementia (n=9, 14.5%).

### 3.5. Demographic, socioeconomic, lifestyle, and comorbidity factors

Over half of the articles investigated demographic influences on ADRD (n=35, 56.5%). Many of these articles identified increasing age as a risk factor for ADRD development (n=15, 42.9%). The impact of sex/gender on ADRD prevalence was consistent with many articles reporting a higher prevalence of dementia in women (n=12, 34.3%), and only two articles finding no significant relationship between sex/gender and dementia (n=2, 5.7%) [51, 59]. One article examined different forms of dementia and found that AD was more prevalent in women, whereas vascular dementia was more common in men (n=1, 2.8%) [69]. Findings on mortality rates by sex were inconclusive— two reported higher mortality rates in women (n=2, 5.7%) [42, 46], and two others reported higher rates in men (n=2, 5.7%) [33, 45]. Additionally, no consistent patterns were observed regarding the effect of race on ADRD. Another notable finding was a higher frequency of diagnostic workups among men (n=1, 2.7%) [52]. Several articles stratified their samples by rural and urban areas to examine demographic influences on ADRD-related health outcomes (n=5, 14.3%). Among those, one article found the highest mortality rates among Asians living in large metropolitan areas, Black individuals in rural areas, and Hispanic individuals in medium metropolitan areas (n=1, 2.8%) [16]. Another article also reported that women in rural areas had a higher prevalence of ADRD, a pattern not observed in urban areas (n=1, 2.8%) [57].

Nearly half of the articles investigated the influence of socioeconomic factors on ADRD (n=30, 48.4%). Among these articles, lower levels of educational attainment (n=20, 66.7%), illiteracy (n=5, 16.7%), low household incomes (n=7, 23.3%), and being single, divorced or widowed (n=3, 10.0%) were consistently identified as risk factors for ADRD [34, 59, 73]. Five articles stratified socioeconomic factors by rural and urban status, identifying education and illiteracy as risk factors in rural areas (n=5, 100%). Among these articles, the majority also recognized education and illiteracy as risk factors in urban areas (n=3, 60%) [21, 22, 59].

Lifestyle factors were examined in a few articles (n=9, 14.5%). Among these articles, inconsistencies were found, with both non-smoking (n=3, 33.3%) [34, 39, 44] and smoking (n=1, 11.1%) [40] identified as risk factors for ADRD. Similarly, physical inactivity (n=2, 22.2%) [38, 57] and physical activity (n=1, 11.1%) [44] were also reported as risk factors. Other findings suggested that activities such as reading and attending social events may protect against ADRD (n=2, 22.2%) [57, 62].

The impact of comorbidities on ADRD was investigated in 16 articles. The presence of 1 to 3 chronic comorbidities increased a person’s risk of ADRD (n=4, 25.0%). Further investigation into specific comorbidities revealed that diabetes (n=6, 37.5%), history of stroke (n=5, 31.3%), cerebrovascular conditions (n=4, 25.0%), hypertension (n=3, 18.8%), and depression (n=2, 12.5%) were identified as risk factors for ADRD.

### 3.6. Environmental factors

Surprisingly, only a small percentage of articles investigated environmental factors and their influence on ADRD (n=8, 12.9%). Among these articles, one article found that for each 5 µg/m³ increase in the air particulate matter, the incidence of ADRD increased by 65% (n=1, 12.5%) [12]. Two articles found that more playgrounds, sports venues, and recreational facilities are associated with decreased ADRD prevalence (n=2, 25.0%) [12, 56]. One article found that individuals with ADRD who live in areas with shortages of mental health professionals are more likely to experience preventable ED visits compared to those who do not live in these areas (n=1, 12.5%) [29]. Moreover, they found that the residents of the most deprived areas experience a significantly lower quality of life score even after controlling for socioeconomic status and comorbidities (n=1, 12.5%) [71]. Only one article stratified results by rural and urban areas and found that individuals from both areas are at a higher risk of ADRD if they work in communities with more blue-collar jobs than those with more professional-class jobs (n=1, 12.5%) [66].

### 3.7. Access, utilization, and quality of care

Over half of all articles (n=32, 51.6%) reported on potential disparities in healthcare access, utilization, or quality. Among these, rural populations were consistently shown to be underdiagnosed, experienced delays in diagnosis, or lacked professional treatment for ADRD compared to their urban counterparts (n=9, 28.1%), with only one such article reporting no difference (n=1, 3.1%) [14]. Additionally, rural residents had fewer encounters with cognitive specialists and mental health providers (n=5, 15.6%). Findings on access and utilization of primary care services were mixed, with an equal number of articles reporting more use (n=2, 6.3%) [52, 74] or less use (n=2, 6.3%) [47, 62] for rural patients. Regarding acute care, rural populations almost always had higher rates of ED visits (n=4, 12.5%), with only one article reporting the opposite (n=1, 3.1%) [50]. Hospitalization rates showed inconsistencies, with two articles reporting higher rates in urban regions (n=2, 6.3%) [27, 62], one article reporting higher rates in rural areas (n=1, 3.1%) [47], and one article finding no significant differences (n=1, 3.1%) [28].

Several articles examined long-term care (LTC) service use (n=8, 25%). Within this subset, consistent findings included rural residents being more likely to be institutionalized in their last year of life (n=2, 25%) [25, 26] and to die in nursing homes (n=2, 25%) [15, 50]. In contrast, urban residents were more likely to access hospice (n=2, 25%) [17, 26] and home health care services (n=2, 25%) [17, 26], and to die in either a hospital, hospice, or home (n=1, 12.5%) [15]. Adjusted mortality and survival outcomes, as proxies for the quality of LTC, were analyzed in a handful of articles (n=8, 25%) that mostly overlapped with those investigating LTC service use. Most articles found rural populations had higher mortality (n=4, 12.5%) [33, 42, 45, 53]. Another found higher urban mortality (n=1, 12.5%) [27], while one article found no difference (n=1, 12.5%) [47]. Survival outcomes reported in these articles also favored urban residents, with (n=2, 25%) articles reporting more prolonged survival for urban participants [25, 26] and one reporting no difference (n=1, 12.5%) [53]. Other quality indicators, such as 30-day readmissions and inappropriate medication prescribing, generally appeared worse for rural groups, though there were exceptions. For example, one study found that potentially preventable hospitalizations were more common among urban people with ADRD (n=1, 12.5%) [47].

### 3.8. Statistical methods used

A variety of statistical methods were employed across the articles. The most used technique was logistic regression (n=23, 37.1%) to assess associations between factors like demographics, healthcare utilization, and ADRD outcomes, often incorporating interaction terms to examine the combined effects of variables. Cox proportional hazards models were also utilized (n=9, 14.5%) to explore time-to-event outcomes such as survival or time to diagnosis, with some articles complementing these analyses using Kaplan-Meier curves (n=4, 6.5%). Poisson regression was employed in a subset of articles (n=6, 9.7%) to estimate prevalence and incidence ratios, adjusting for multiple covariates. Negative binomial regression (n=3, 4.8%) was used to model count data such as symptom occurrence and mortality [13, 45, 72]. Often, these regressions informed multivariate and multilevel models used in many studies (n=10,16.1%), controlling multiple confounding factors while accounting for nested data structures, such as clustering by region or healthcare facility.

## 4. DISCUSSION

To our knowledge, this is the first study that provides a comprehensive overview of the current worldwide state of rural-urban disparities in ADRD, focusing on identifying key associated risk factors and highlighting gaps in various aspects of disparities. Following PRISMA guidelines, we identified 62 articles that investigated rural-urban disparities in ADRD with no restrictions on geography or study design. Most articles were published after 2010, particularly within the past 6 years. A substantial portion of the articles were conducted in the US, China, and Canada. Most articles had cross-sectional or cohort study designs. The main outcomes were the prevalence or incidence of ADRD, though several articles also examined mortality and healthcare utilization. Rural-urban classification systems were inconsistent, leading to a lack of a clear definition of rurality across articles. Notably, only a small percentage of articles investigated the role of environmental or lifestyle factors and their influence on disparities, representing a significant gap in current literature. The review offers a foundation for sustained research on rural-urban disparities in ADRD and encourages researchers to explore the identified gaps and foster innovative approaches for future investigations in various areas of rural-urban disparities of ADRD.

A previous systematic review explored geographical variation in dementia across cross-sectional and longitudinal study designs worldwide [10]. They examined how different diagnostic criteria affected the relationship between rural residence and dementia prevalence. They found that patients in rural communities experienced higher mortality rates and worse healthcare access. Additionally, the use of the DSM-IV (Diagnostic and Statistical Manual of Mental Disorders, Fourth Edition) or the 10/66 criteria (a framework for diagnosing dementia in developing countries) was shown to significantly influence these outcomes. However, their findings were limited by inconsistent definitions of rurality, variation in countries of origin, and the absence of an investigation into other risk factors beyond rural residence. An earlier systematic review focused on disparities in healthcare quality among dementia patients in rural and urban areas [9]. Their results concerning mortality and medical care utilization were consistent with our review. They also found a higher prescription rate of anti-dementia medications in rural areas compared to urban ones. However, this review did not report on potential factors contributing to these disparities and queried only one online database to identify eligible articles.

Most included studies were conducted in the US, China, and Canada. Several factors likely contribute to the concentration of research in these countries, including an aging population, investment in research, and a focus on relevant policies. All three countries are experiencing rapidly aging populations [75]. Since age is a major risk factor for chronic diseases like ADRD, these countries face a greater need for research and investment in ADRD, particularly in identifying risk factors and understanding the underlying mechanisms. Additionally, these countries have well-developed healthcare and research infrastructures supported by significant investments in health research [76]. Conversely, very few articles have been conducted in developing and underdeveloped countries. An international team led by the Alzheimer’s Association published a report on rural health disparities in ADRD, emphasizing that undiagnosed dementia, lack of resources, and economic challenges are most severe in low- and middle-income countries [77]. However, the evidence, especially derived from ADRD studies in developing countries, to support and enhance these calls for collective action remains sparse. Many developing countries have limited healthcare budgets and prioritize addressing communicable diseases [78]. To gain a more comprehensive understanding of ADRD’s global impact, future studies should prioritize research in underrepresented regions.

The most common demographic risk factors for ADRD identified in all included studies were age and being female. Age is a well-established risk factor for ADRD, likely because the brain undergoes natural changes over time (such as changes to brain cells, proteins or plaques, reduced brain plasticity, and cumulative damage from lifestyle factors), some of which increase the risk of cognitive decline [79]. Recent research advancements have identified the neuroprotective properties of estrogen in females in relation to neurodegenerative diseases and brain injuries [80]. The decline in estrogen levels that women experience after menopause may contribute to their higher risk of developing ADRD. Additionally, women tend to live longer than men, contributing to a higher overall exposure to aging-related risk factors for ADRD, thereby increasing the prevalence of ADRD among women. Among the socioeconomic variables, low educational attainment was the most common risk factor. Individuals with lower education levels often earn lower wages, which may limit their ability to afford healthcare, nutritious food, and safe living conditions, leading to poorer health outcomes compared to those with higher levels of education [81]. While the results for lifestyle factors investigated in these articles were mostly unclear, physical inactivity was found to be a more common risk factor for ADRD. Physical activity has long been recognized as a key component of a healthy life that improves blood flow to the brain and reduces inflammation [82].

Very few articles have investigated environmental influences on ADRD. However, among those that have, a higher number of playgrounds, community centers, and sports venues within a community was associated with a decreased prevalence of ADRD. This finding may be explained by the increased likelihood of physical activity in communities with more recreational facilities, which can lower ADRD risk by promoting blood flow to the brain and reducing inflammation [82]. Additionally, one article found that greater exposure to air pollution increases a person’s risk of developing ADRD. This relationship may stem from inhaled gasses, particles, or substances desorbed from the particle surface, which can induce inflammatory responses, activate microglia, and increase the production of Aβ peptides, possibly related to cognitive decline [83].

Among the various comorbidities examined, diabetes, cerebrovascular conditions, and hypertension were the most reported. While the mechanisms are not yet fully understood, it is well established that diabetes is associated with changes in cognition, including learning, memory, mental speed, and mental flexibility [84]. These changes may result from diabetes affecting the brain’s ability to use glucose, its primary energy source [84]. Additionally, cerebrovascular diseases often reduce blood flow to the brain, increasing cell damage and cognitive impairment [85]. Hypertension is known to damage blood vessels, leading to decreased blood flow to the brain and increased white matter lesions and atrophy, which are associated with cognitive impairment and ADRD [86].

Most included studies employed traditional statistical methods such as logistic regression, Cox proportional hazards, Poisson regression, and negative binomial regression. These methods were typically embedded in multivariate models and adjusted for common baseline demographic factors, such as age, sex, race/ethnicity, and, to a lesser extent, education, occupation, marital status, and comorbidities. Thus, these studies controlled for individual-level factors routinely captured in large, often national, administrative datasets. However, few studies were able to adjust for less commonly available individual or group-level variables, such as functional status, genetic predisposition, or environmental factors. Such factors are often highly influential but context-specific and thus challenging to capture on a large scale using billing claims or standardized surveys [74, 87, 88]. Similarly, most studies did not account for area-level deprivation (e.g., crime, pollution, and segregation), enrichment (e.g., green space, public transportation, and community inclusion), and key social determinants of health. These contextual factors, especially at the smaller geographic scales such as neighborhoods or ZIP codes, may play critical roles in shaping ADRD disparities [6, 89–92]. Furthermore, while some advanced statistical techniques, such as Bayesian models, address spatial variability, the limited granularity of these analyses can still overlook finer-scale geographic influences [12]. Machine learning (ML) and large language models (LLMs) represent a promising means of addressing some of these limitations. By applying ML techniques to large datasets, the identification and differentiation of people with ADRD by type or severity, which is currently hampered by inadequate coding and widespread underdiagnosis, is steadily improving [93, 94]. LLMs, which can process vast amounts of unstructured text data, may also offer new ways to glean insights from clinical notes or other narrative data sources that have historically been difficult to analyze [95].

In reviewing other methodological limitations across the included articles, several recurring themes were identified. A key issue was the heavy reliance on secondary data sources, such as large administrative or claims datasets, which raised concerns about the accuracy of data and the absence of individual-level data on dementia type and severity. The challenge of detecting dementia, both in non-clinical and, to a lesser extent, clinical settings, contributed to a near-universal problem of dementia misclassification. Lack of generalizability of results across dementia treatment populations was another common limitation, particularly in articles focusing on specific and distinct subpopulations, such as Medicare Fee-For-Service beneficiaries [17, 25], Veterans [96], or residents of smaller geographic areas (e.g., single-county analyses). Several articles pointed out that the simplistic categorization of rural vs. urban using area-level indicators (i.e., county or even census tract) fails to capture the full complexity and heterogeneity within these regions. Many included articles were cross-sectional designs, limiting their ability to draw causal inferences or capture temporal changes in ADRD outcomes and healthcare use. This is a significant gap, as longitudinal analyses would allow for a better understanding of how risk factors and interventions influence dementia outcomes over time. Selection bias was commonly highlighted, especially in articles that excluded institutionalized, had age or literacy requirements, relied on self-reported or proxy-reported data, or reported lower participation rates and sample sizes in rural compared to urban groups, compromising the reliability of inferences. The possibility of relocation bias, which can arise if individuals who moved between rural and urban areas differed systematically from those who remained in place, was raised by some authors. These limitations highlight a clear need for more rigorous perspectives to strengthen the evidence.

There are several limitations to this review. First, we included only articles published in English, which may have led to the exclusion of important articles from non-English regions. As a result, our findings predominantly reflect the English-speaking world or regions with resources for translation. Moreover, the variety of rural-urban classification systems complicates the comparison of article results. While one article may report significant findings based on its specific classification, those results might not hold under different systems. We recommend that future studies explore causality or mediation analysis rather than mere associations to understand the mechanism shaping these disparities. A meta-analysis would be valuable for pooling the articles of the same design or those employing identical rural-urban classification systems, thereby better addressing the limitations identified in this review. Additionally, future research could focus more on variables that have not been deeply explored here, such as the genetic factors or the effectiveness of governmental interventions and policies.

## 5. CONCLUSION

This scoping review provided a comprehensive overview of the current worldwide state of rural-urban disparities in ADRD, focusing on identifying key associated risk factors and highlighting gaps in various aspects of disparities. A dearth of novel and sophisticated approaches, especially related to rural-urban classification systems as well as ADRD detection and differentiation, suggests opportunities for innovative research that employs advanced or emerging methodologies. Most prior studies have focused on a limited set of demographic indicators due to data source constraints, leaving room to explore other influential but under-investigated risk factors for ADRD. Finally, rigorous and robust longitudinal analyses, which could more accurately capture the underlying causes and track the progression of ADRD, are needed.

## Data Availability

All data produced in the present work are contained in the manuscript

